# Pulsatile Brain Motion as a Marker of Brain Aging and Dementia: Insights from 3D q-aMRI

**DOI:** 10.64898/2025.12.10.25341649

**Authors:** Itamar Terem, Kyan Younes, Skylar Weiss, Andrew Dreisbach, Hillary Vossler, Eryn Kwon, Daniel Cornfeld, Jet Wright, Paul Condron, Kathleen L Poston, Victor W. Henderson, Elizabeth C Mormino, Samantha Holdsworth, Kawin Setsompop

## Abstract

Heart-brain interactions, including cardiac-induced brain pulsatility, are thought to support brain homeostasis, yet their alteration with aging and neurodegeneration remains poorly understood. Here, we used three-dimensional quantitative amplified MRI (q-aMRI) to visualize and quantify cardiac-induced pulsatile brain motion across healthy individuals and those on the Alzheimer’s and Lewy body disease spectra. Expert evaluations revealed consistent distinctions between normal and abnormal motion patterns, which were further characterized using strain-based biomechanical features. Principal component analysis identified interpretable signatures of abnormal motion that emerged predominantly after midlife and were closely linked to chronological age. Even after accounting for age, these biomechanical features differentiated individuals with clinical and biomarker evidence of neurodegenerative disease. Together, these findings suggest that q-aMRI–derived measures of cardiac-gated pulsatile brain motion may serve as imaging biomarkers of normal brain aging and dementia.

## Introduction

With each cardiac pulsation, dynamic fluctuations in blood vessels and cerebrospinal fluid (CSF) flow induce periodic deformation of brain tissue^1–6^. This intrinsic pulsatile brain motion is governed by tissue mechanical properties, vascular compliance, intracranial pressure, and flow dynamics^5,7–9^. Understanding this mechanical behavior is critical for elucidating key physiological processes and detecting early pathological changes.

Recent research has highlighted cardiac-induced pulsatility as a potential vital driver of brain homeostasis. CSF circulation^10^, glymphatic clearance^11^, and perivascular transport^12^ may depend on pulsatile motion and are sensitive to alterations in brain compliance, vascular health, and age-related structural remodeling^13^. Aging disrupts brain biomechanics via arterial stiffening^14^, reduced tissue elasticity, and impaired CSF dynamics^15,16^ compromising metabolic waste clearance and potentially contributing to cognitive decline and neurodegenerative disorders such as Alzheimer’s disease (AD)^17^. The brain’s diminished capacity to accommodate pulsatile mechanical input during aging may disturb the balance between tissue motion and fluid transport, suggesting brain motion characteristics as potential biomarkers of healthy aging and early disease.

AD and Lewy body disease (LBD) represent the two most common neurodegenerative disorders^18^. Increasing evidence demonstrates that these pathologies frequently co-occur, making polypathology the rule rather than the exception^19–22^. The high prevalence of mixed pathology underscores the need to elucidate shared upstream mechanisms and to develop biomarkers that provide mechanistic insights rather than serving solely diagnostic purposes^23^. Contemporary diagnostic criteria for AD^24^ and LBD^25^ incorporate biological staging based on biomarker profiles, rather than relying exclusively on clinical impairment. Consequently, individuals meeting criteria for AD or LBD may exhibit a spectrum of clinical presentations, ranging from cognitively unimpaired to mild cognitive impairment (MCI) or dementia. This gradation in staging offers an opportunity to interrogate early mechanistic alterations that may predispose to or drive the development of co-pathologies.

Amplified Magnetic Resonance Imaging (aMRI^26–30^) is an emerging technique that enables the visualization of pulsatile brain motion by amplifying subtle sub-voxel motion present but unseen in cardiac-gated cine MRI data. This technique produces high-contrast, high SNR, high-temporal-resolution videos that reveal the brain’s pulsatile motion with great details. This approach has shown promise in studying neurological disorders^31–35^ and evaluating CSF computational fluid dynamics^36^. However, conventional 3D aMRI provides only qualitative visualization without direct measurement of tissue displacement.

This limitation has recently been addressed with the development of three-dimensional quantitative aMRI^37^ (3D q-aMRI) – an extension of 3D aMRI that enables voxel-level quantification of the displacement field. With specialized post-processing, 3D q-aMRI enables visualization and precise quantification of sub-voxel tissue motion, providing a novel, noninvasive window into brain biomechanics and showing early promise for identifying abnormal motion patterns in neurodegenerative disease^37^.

In this study, we applied 3D q-aMRI to a cohort of adults spanning the full adult lifespan, including cognitively healthy individuals and participants along the AD and LBD spectra. Our goals were to characterize age-related alterations in cardiac-induced brain motion and to determine whether deviations from normal motion patterns are associated with neurodegenerative disease. To this end, expert raters evaluated amplified brain motion videos to establish normative and abnormal patterns, while biomechanical strain features were derived from the underlying displacement fields to capture regional brain tissue deformation. Data-driven analyses using principal component decomposition and regression modeling were used to identify dominant motion signatures, their dependence on aging, and their associations with clinical and AD biomarker status.

## Method

### Human Participants

Ethical approval was obtained from the Stanford University Institutional Review Board and the University of Auckland Human Participants Ethics Committee. Written informed consent was obtained from all participants. Healthy aging participants and participants with neurodegenerative diseases were recruited from the Stanford Alzheimer’s Disease Research Center and clinical diagnosis was made per published criteria^24,25^ based on a multidisciplinary evaluation and a consensus conference that included behavioral neurology, neuropsychology, and nursing input. A separate cohort of healthy participants younger than 50, originally collected during the development of 3D q-aMRI, was also included. Data were collected from 147 participants, of whom 105 passed quality control and were included in the analysis. Quality control involved two steps. First, raw data were visually inspected, and datasets with significant involuntary subject head motion, blurring, or excessive noise were excluded. Second, amplified videos were reviewed, and participants exhibiting apparent skull motion –an artifact not expected in stable anatomy –were also excluded. After quality control, 105 participants (55 females, 50 males; age range 18–93 years, median ± IQR: 69.98 ± 21.91 years) were included in the final analysis (Table 1). 31 (∼30%) participants were amyloid-PET positive, 47 (∼45%) were amyloid-PET negative, and 27 (∼25%) had unknown amyloid-PET status. Notably, most individuals with unknown amyloid-PET status were drawn from the younger cohort.

**Table 1.**
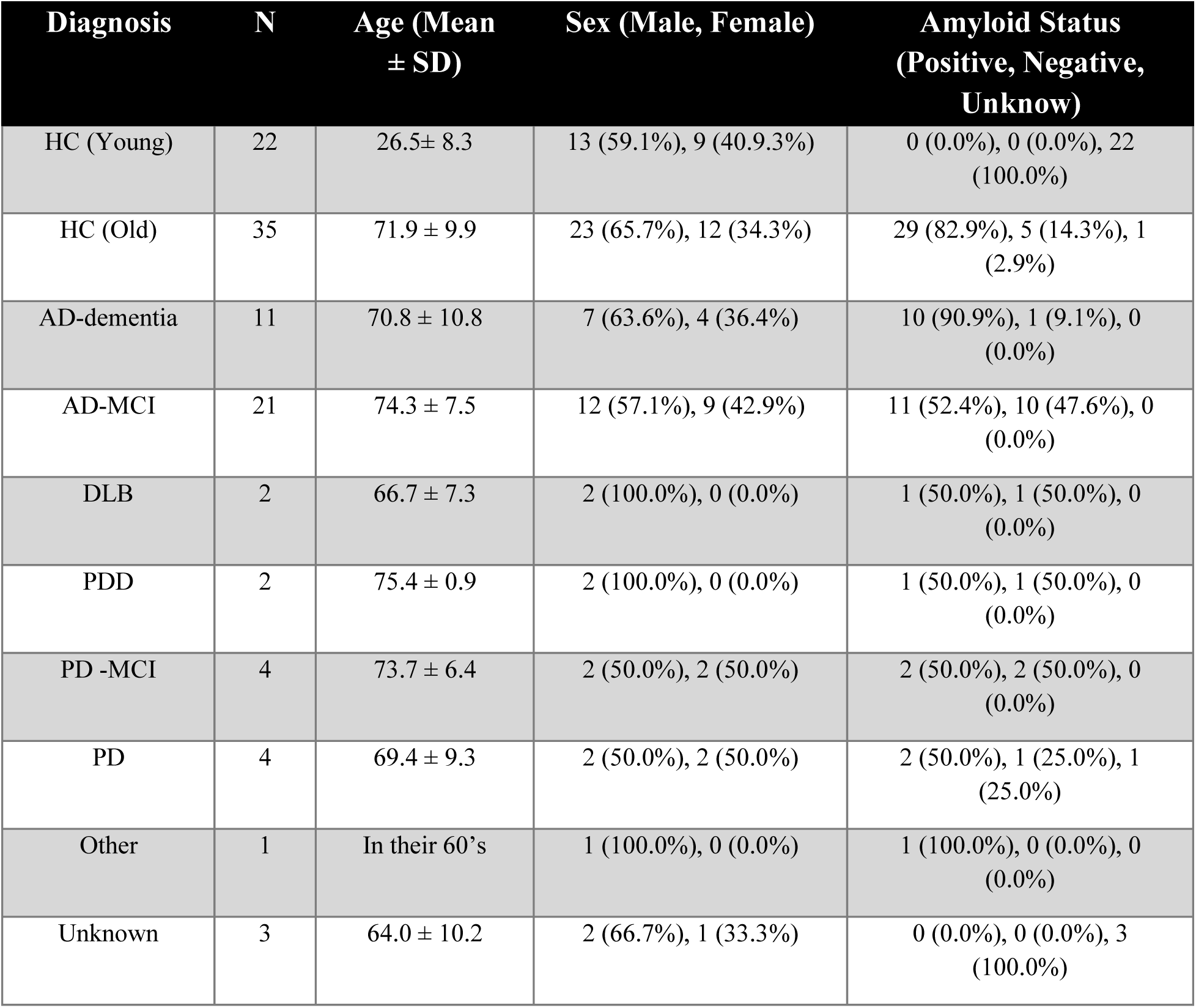
Cohort demographics by diagnostic group, including number of participants, sex, age, and amyloid status. Abbreviations: HC – Healthy Control; AD – Alzheimer’s disease; MCI – Mild Cognitive Impairment; DLB – Dementia with Lewy bodies; PD – Parkinson’s disease; PDD - Parkinson’s disease dementia; Other – non-AD neurological conditions (corticobasal syndrome, semantic variant primary progressive aphasia, PTSD).

| Diagnosis | N | Age (Mean<br>± SD) | Sex (Male, Female) | Amyloid Status<br>(Positive, Negative,<br>Unknow) |
| --- | --- | --- | --- | --- |
| HC (Young) | 22 | 26.5± 8.3 | 13 (59.1%), 9 (40.9.3%) | 0 (0.0%), 0 (0.0%), 22 (100.0%) |
| HC (Old) | 35 | 71.9 ± 9.9 | 23 (65.7%), 12 (34.3%) | 29 (82.9%), 5 (14.3%), 1 (2.9%) |
| AD-dementia | 11 | 70.8 ± 10.8 | 7 (63.6%), 4 (36.4%) | 10 (90.9%), 1 (9.1%), 0 (0.0%) |
| AD-MCI | 21 | 74.3 ± 7.5 | 12 (57.1%), 9 (42.9%) | 11 (52.4%), 10 (47.6%), 0 (0.0%) |
| DLB | 2 | 66.7 ± 7.3 | 2 (100.0%), 0 (0.0%) | 1 (50.0%), 1 (50.0%), 0 (0.0%) |
| PDD | 2 | 75.4 ± 0.9 | 2 (100.0%), 0 (0.0%) | 1 (50.0%), 1 (50.0%), 0 (0.0%) |
| PD -MCI | 4 | 73.7 ± 6.4 | 2 (50.0%), 2 (50.0%) | 2 (50.0%), 2 (50.0%), 0 (0.0%) |
| PD | 4 | 69.4 ± 9.3 | 2 (50.0%), 2 (50.0%) | 2 (50.0%), 1 (25.0%), 1 (25.0%) |
| Other | 1 | In their 60's | 1 (100.0%), 0 (0.0%) | 1 (100.0%), 0 (0.0%), 0 (0.0%) |
| Unknown | 3 | 64.0 ± 10.2 | 2 (66.7%), 1 (33.3%) | 0 (0.0%), 0 (0.0%), 3 (100.0%) |

In terms of clinical classification, 58 participants were cognitively normal controls, 3 had an unknown diagnosis, and 44 had a clinical diagnosis: 11 with dementia due to AD, 21 with MCI due to AD, 2 with dementia due to LBD, 3 with Parkinson’s disease (PD) without cognitive impairment, 4 with PD with MCI, 2 with PD dementia, and 1 with another non–AD-related neurological condition.

### MRI acquisition

Data were acquired at two sites: Stanford University (96 participants) and the Mātai Medical Research Institute in New Zealand (9 participants), using three different scanners. Two of the scanners were 3T MRI systems (SIGNA Premier; GE Healthcare, Milwaukee, WI, USA), and the third was a hybrid PET/MR scanner (also SIGNA Premier; GE Healthcare).

All scanners used the same standardized 3D q-aMRI acquisition protocol: whole-brain, 3D volumetric, cardiac-gated cine MRI datasets were collected in the sagittal plane using a balanced steady-state free precession (bSSFP/FIESTA) sequence with the following parameters: field of view (FOV) = 24 × 24 cm², matrix size = 200 × 200, TR/TE/flip angle = 2.8 ms / 1.4 ms / 25°, acceleration factor = 3 or 8, spatial resolution = 1.2 mm isotropic, and peripheral pulse gating with retrospective binning into 20 cardiac phases. A total of 120 slices were acquired to ensure full brain coverage. Scan times were approximately 2 minutes and 30 seconds with an acceleration factor of 8, and around 6 minutes with an acceleration factor of 3.

PET/MR scans were performed only on the disease cohort (81 participants) using a simultaneous time-of-flight (TOF)-enabled PET/MR scanner with high sensitivity (23.3 cps/kBq). MRI data were collected simultaneously with PET imaging. High-resolution T1-weighted spoiled gradient recalled echo (SPGR) scans (TR/TE/TI = 7.7 / 3.1 / 400 ms, flip angle = 11°, resolution = 1.2 × 1.1 × 1.1 mm³) were acquired and used to define regions of interest (ROIs) in each participant’s native space using the Free Surfer software package^38^ (version 6.0; Fischl, 2012; http://surfer.nmr.mgh.harvard.edu/). Amyloid radiotracer F-florbetaben (Life Molecular Imaging, Berlin, Germany) was injected intravenously and PET data were acquired 90–110 min after injection. Frames were motion-corrected and realigned to the T1-weighted image, then summed to generate a single image for each scan. Amyloid status was determined by normalizing the summed PET signal using the mean uptake in the whole inferior cerebellar gray matter as a reference region to extract global standardized uptake ration (SUVR)^39^.

### Data Processing

For each participant, the following analyses were performed (Fig.1a). 3D q-aMRI was applied to generate both amplified motion videos and voxel-wise displacement fields. 3D q-aMRI is a post-processing technique that visualizes and quantifies subtle, pulsatile brain tissue motion that is typically imperceptible in standard cardiac-gated cine MRI. The method decomposes the 3D cine MRI data using a multiscale, multi-orientation steerable pyramid transform, capturing motion across a range of spatial frequencies and orientations. Motion sensitivity is achieved by manipulating the phase component of the complex-valued signal, which encodes local displacements over time, enabling detection of minute movements while preserving anatomical fidelity^37^.

The algorithm operates via two complementary pathways: (1) a visualization pathway, which amplifies motion to produce intuitive, exaggerated videos of brain movement, and (2) a quantification pathway, which computes voxel-wise displacement fields across time by solving the optical flow equation over the filtered phases, enabling downstream biomechanical and statistical analysis. For all analyses, hyperparameters were set as follows: magnification factor = 30, band-pass filter centered on the first cardiac harmonic, and spatial smoothing (σ) = 5.

The raw average cine bSSFP volume was used to compute the rigid-body transformation to MNI space in MATLAB (MathWorks, Natick, MA, USA, 2024). Regions of interest (ROIs) included the ventricular system, cerebral cortex and white matter, thalamus, basal ganglia (caudate, putamen, pallidum), hippocampus, amygdala, accumbens area, ventral diencephalon, and brainstem were segmented^40^ for region-based analysis. These regions were selected to capture midline structures and key subcortical areas implicated in neurodegenerative diseases: the basal ganglia are relevant to Lewy body disease and Parkinson’s disease, the hippocampus is critically involved in Alzheimer’s disease, and the ventricular system and other midline structures have been shown to undergo measurable brain motion^41^. The bSSFP acquisition is highly sensitive to magnetic field inhomogeneities, which can produce image artifacts^42^ that propagate as errors in sub-voxel displacement field estimation by 3D q-aMRI. As a result, the following regions were excluded from the final analysis: brainstem, ventral diencephalon, and nucleus accumbens.

Amplified videos were used for expert consensus evaluation and ground-truth labeling, while the voxel displacement fields were used to compute strain tensors at each voxel. The strain tensor characterizes local tissue deformation in three dimensions over the cardiac cycle, capturing both compressive and shear motion^43^. From these strain tensors, we derived multiple biomechanical measures to characterize tissue deformation. Volumetric strain quantifies local tissue expansion or compression, while octahedral shear strain captures shear deformation independent of volumetric changes. Strain energy reflects the mechanical energy stored in the tissue during deformation. Eigenvalues represent the magnitudes of the principal strains, indicating the intensity of deformation along the principal axes. The corresponding principal vectors indicate the orientation of the maximum and minimum strain, providing directional information. Together, these measures allow a comprehensive characterization of both the magnitude and orientation of biomechanical changes across brain regions and cardiac phases.

Strain tensor–derived biomechanical measures were computed within each ROI. Except for the principal vectors, all measures are invariant to rigid transformations, so only the principal vectors were registered to MNI space to align their directional components (X, Y, Z) with standard anatomical axes – superior–inferior (SI), anterior–posterior (AP), and left–right (LR). For each cardiac phase, we quantified the distribution of these measures within each ROI using seven statistical descriptors (mean, variance, median, mode, skewness, kurtosis, entropy). The resulting features were concatenated into a single vector of length 51,870 (26 ROIs × 15 biomechanical measures × 7 statistical descriptors × 19 cardiac phases), producing a data matrix of 105 participants × 51,870 features.

### Reader-Based Consensus Labeling

For each subject, twelve amplified videos were generated: six axial-plane views at the level of the lateral ventricles and six sagittal-plane views centered on the midsagittal slice. Five readers participated, including two aMRI experts (I.T., S.H.), two with moderate experience (E.K., J.W.), and one radiologist with limited prior exposure to aMRI (D.C.). Readers were blinded to participant characteristics and independently assessed the presence or absence of three predefined motion features, empirically determined by I.T. based on extensive experience with aMRI data: each dataset was reviewed individually, recurring patterns of brain motion were identified, and features in key brain regions were selected as most informative for distinguishing normal from abnormal motion.

1. Lateral ventricles (axial slice): Symmetrical, well-synchronized expansion and compression.
2. Fourth ventricle (sagittal slice): Predominantly superior–inferior motion with minimal anterior–posterior displacement.
3. Synchronization (sagittal slice): SI-directed motion of ventricular and midline structures with minimal phase delay.

A subject was classified as abnormal if any feature was absent. Readers could also annotate ambiguous cases or additional observations. Intra-reader reliability was assessed using nine participants (8.6%) presented twice in a blinded manner. Inter-reader agreement was quantified using Fleiss’ kappa^44^, measuring consistency beyond chance. Final consensus labels were established in a structured meeting, where all disagreements were discussed and resolved collaboratively.

### Principal Component Analysis (PCA) Features Analysis

The feature data matrix comprised 105 participants and 51,870 features. Given the high dimensionality relative to the sample size, PCA was applied to reduce dimensionality, mitigate overfitting, and identify features contributing most to differences between reader-classified normal and abnormal motion patterns.

Group separation in PCA space was quantified using the absolute value of Cohen’s d for each principal component (PC). Cohen’s d expresses the standardized difference between group means, providing a scale-independent measure of separation. PCs with |d| ≥ 0.2 – the conventional threshold^45^ for a small but meaningful effect size – were retained. Feature-level contributions were computed by weighting absolute PCA loadings by the corresponding Cohen’s d values, such that feature importance reflected both PCA structure and the degree of group separation. To assess statistical significance, group labels were permuted 1,000 times, and contributions were recomputed to form an empirical null distribution. Two-tailed empirical p-values were obtained as the fraction of permuted contributions more extreme than the observed value. Multiple comparisons were controlled using the false discovery rate (FDR, q < 0.05). Features surviving FDR correction were aggregated by anatomical region, strain tensor component, statistical descriptor, and cardiac phase. Aggregated values were visualized as grouped contribution matrices summarizing regional–strain, regional–statistical, and regional–temporal relationships.

### Machine Learning-Based Brain Motion Classification

Principal components (PCs) derived from the feature matrix were used as inputs to five predictive models, following the order of analyses presented in the Results section. Unless otherwise noted, only PCs explaining 90% of the variance were retained for model training. To prevent information leakage, PCA was recomputed independently within each training fold for all five models. Model performance was estimated using stratified 4-fold cross-validation repeated 25 times (100 train–test evaluations), providing stable metrics and robust estimates of variability.

#### Model 1: Logistic regression for normal vs. abnormal motion

The first model was a logistic regression classifier trained to distinguish normal from abnormal brain motion. The full dataset was divided into a training set of 79 participants and a test set of 26 participants. L2 regularization and balanced class weights were used to mitigate overfitting and address class imbalance. This model served as a classifier for detecting abnormal motion patterns.

#### Models 2–3: Elastic net regression for brain-age prediction

Models 2, and 3 were all elastic net regression models using identical hyperparameters (regularization strength α = 0.01 and L1 ratio = 0.5). These models differed only in the subset of participants used for training, enabling complementary ‘normative’ aging analyses:

● **Model 2:** Trained on the complete dataset, 79-participant training set and evaluated on 26-participant test set.
● **Model 3:** Trained exclusively on participants with normal brain motion (n = 49). Participants with abnormal motion (n = 56) were then projected into this model to quantify deviations from the expected motion-based aging trajectory.

#### Model 4: Logistic regression for adults over 50 years

The final model was a logistic regression classifier trained to distinguish normal from abnormal motion among participants older than 50 years, the age group most relevant to dementia and neurodegeneration. The training set consisted of 58 participants, with 20 held out for testing. To isolate age-independent effects, chronological age was regressed out of the features prior to model training. Because of the smaller sample size, only the first seven PCs were used to reduce overfitting. Average predicted probabilities across the 100 cross-validation iterations were used to assess associations between abnormal motion, clinical diagnosis, and amyloid status.

### Statistical Analysis and Performances Evaluation

Inter-rater agreement among expert readers was assessed using Fleiss’ Kappa, with corresponding 95% confidence intervals to evaluate the reliability and consistency of visual assessments. Confidence intervals (CIs) were computed using the t-distribution based on the standard error of the mean (SEM) to quantify uncertainty in labeling and model performance metrics. For the logistic regression classifier, performance metrics included area under the receiver operating characteristic curve (AUC), sensitivity, specificity, accuracy, and F1 score, all reported with 95% CIs. Additionally, the model’s predicted mean probability for each subject was presented alongside its 95% CI to assess prediction uncertainty. For regression analyses, we reported the mean absolute error (MAE) and the coefficient of determination (R²) with their respective 95% confidence intervals. In addition, we calculated the Pearson correlation coefficient (r) between predicted and chronological age, along with its associated p-value, to assess the strength and significance of the linear association. To test associations between abnormal brain motion, neurodegenerative disease diagnosis, and amyloid PET status, we applied a one-sided Fisher’s exact test, appropriate for categorical data and small sample sizes. All statistical analyses were conducted using Python.

## Results

Of the 42 participants excluded during quality control, the majority were cognitively normal healthy controls (HC, ∼43% of excluded cases), while the remaining participants included individuals with neurodegenerative conditions.

The overall data processing and analysis workflow is illustrated in Fig. 1a. For each participant, cardiac-gated cine MRI data were processed using 3D q-aMRI to generate amplified motion videos and voxel-wise displacement fields. The amplified videos were visually assessed by expert readers to classify brain motion as normal or abnormal, providing reference labels for quantitative analyses. Displacement fields were used to compute strain tensors and derive biomechanical features (Fig. 1b), which were assessed within anatomical regions and across cardiac phases. These region-level biomechanical descriptors formed the input for dimensionality reduction and predictive modeling of motion classification and brain age.

**Fig. 1:**
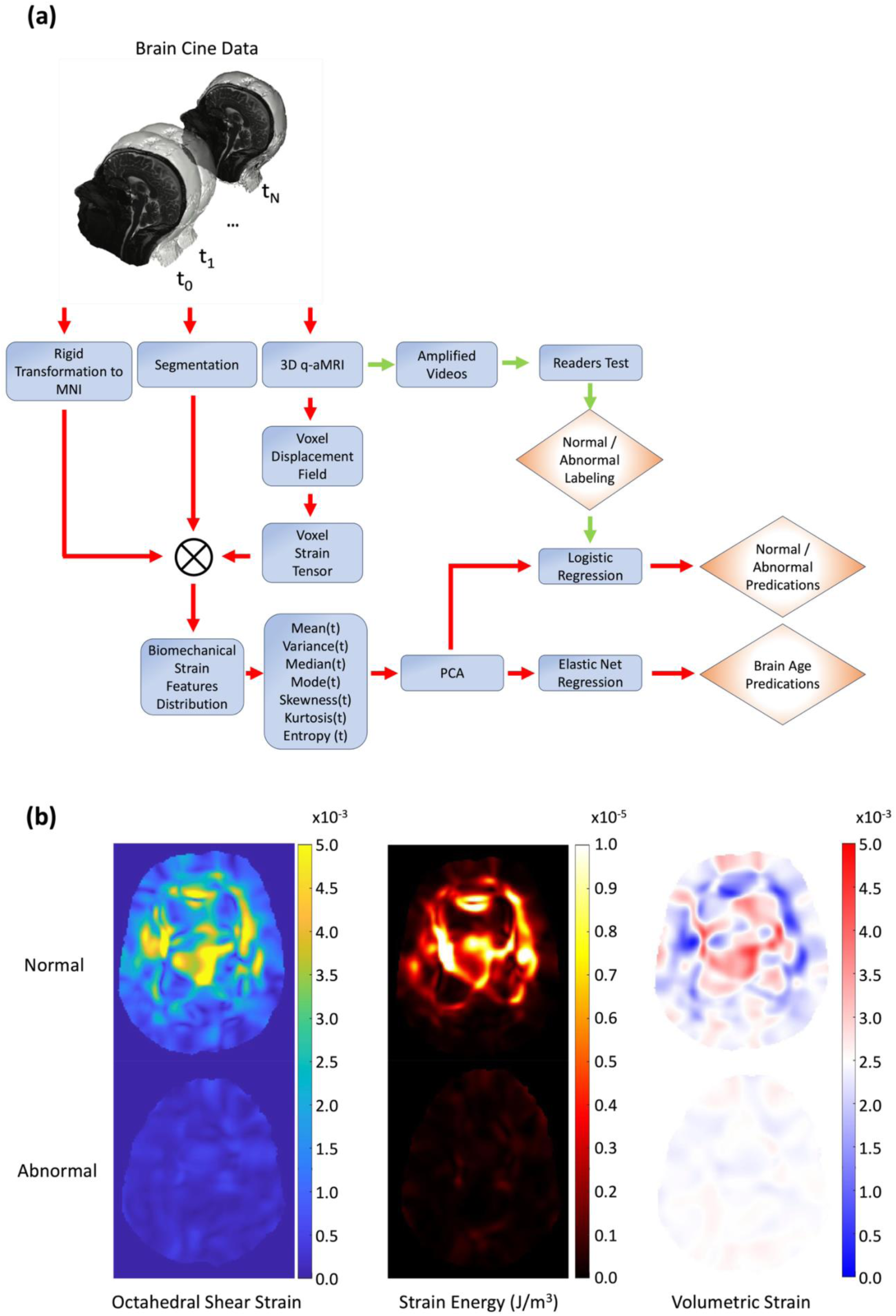
Overview of the data processing pipeline (a) and example biomechanical feature maps (b). For each participant, cardiac-gated cine MRI data were processed using 3D quantitative amplified MRI (3D q-aMRI) to generate amplified motion videos and voxel-wise displacement fields. The amplified videos were used for expert reader labeling to classify brain motion as normal or abnormal. Voxel-wise displacement fields were further used to compute strain tensors and derive biomechanical features. Regions of interest (ROIs) were segmented, and the biomechanical features were registered to MNI space using a rigid-body transformation. For each ROI, time-varying distributions of biomechanical features were computed across 19 cardiac phases, and seven statistical descriptors were extracted per phase, resulting in a feature vector of 51,870 dimensions per participant. Principal component analysis (PCA) was applied to reduce dimensionality, and the resulting components were used to train a logistic regression model for motion classification and an elastic regression model to predict brain age. (b) shows example axial-plane maps at the level of the lateral ventricles of octahedral shear strain and volumetric strain for one subject classified as having normal motion and one subject classified as having abnormal motion. Clear differences are visible, with abnormal motion showing reduced magnitude compared to normal motion.

### Reader-Based Consensus Labeling

Inter-rater reliability among the five readers – two aMRI experts, two researchers with moderate experience, and one radiologist with limited aMRI experience – was substantial, with a Fleiss’ Kappa of 0.665 (standard error = 0.031; 95% CI: 0.649–0.680). The corresponding z-statistic was 21.53 (p < 0.0001), rejecting the null hypothesis that the observed agreement occurred by chance.

Of the 105 participants, 68 cases showed complete agreement, 24 cases had one dissenting reader, and 13 cases were split 3–2. Discrepancies were resolved during a consensus meeting, which also refined the qualitative criteria for differentiating normal from abnormal motion. Specifically, it was agreed that under normal motion, the ventricular system and midline structures should exhibit superior–inferior displacement of comparable magnitude. Incorporating such refinements, together with more targeted reader training, is expected to further enhance consistency in future assessments. Following consensus, 93 cases achieved complete agreement, 7 cases had one dissenting vote, and 5 remained split (2–3).

The final consensus yielded 49 participants with normal motion and 56 with abnormal motion, which served as the reference standard for subsequent quantitative analyses of brain motion features. Intra-reader reliability, assessed on nine duplicate cases (4 normal, 5 abnormal), showed perfect agreement for Readers 1 and 2 (I.T., S.H.; 9/9) and near-perfect agreement for Readers 3–5 (E.K., J.W., D.C.; 8/9).

### PCA Features Analysis

To identify biomechanical features that best differentiate normal from abnormal brain motion, we first performed PCA on the full feature matrix. The degree of group separation along each component was quantified using Cohen’s *d*, and eight principal components exceeded the conventional threshold for a small but meaningful effect size (|*d*| ≥ 0.2). Among these, PC1 and PC2 showed the strongest separation (|*d*| = 1.29 and 1.70, respectively).

Projection of the feature data onto the first two components (Fig. 2a) revealed a clear separation between participants classified as normal and those with abnormal motion patterns based on five-reader consensus. Participants for whom expert readers had difficulty reaching consensus (marked with black circles) clustered near the boundary between normal and abnormal groups, indicating that their motion patterns were also intermediate in the model’s feature space.

**Fig. 2:**
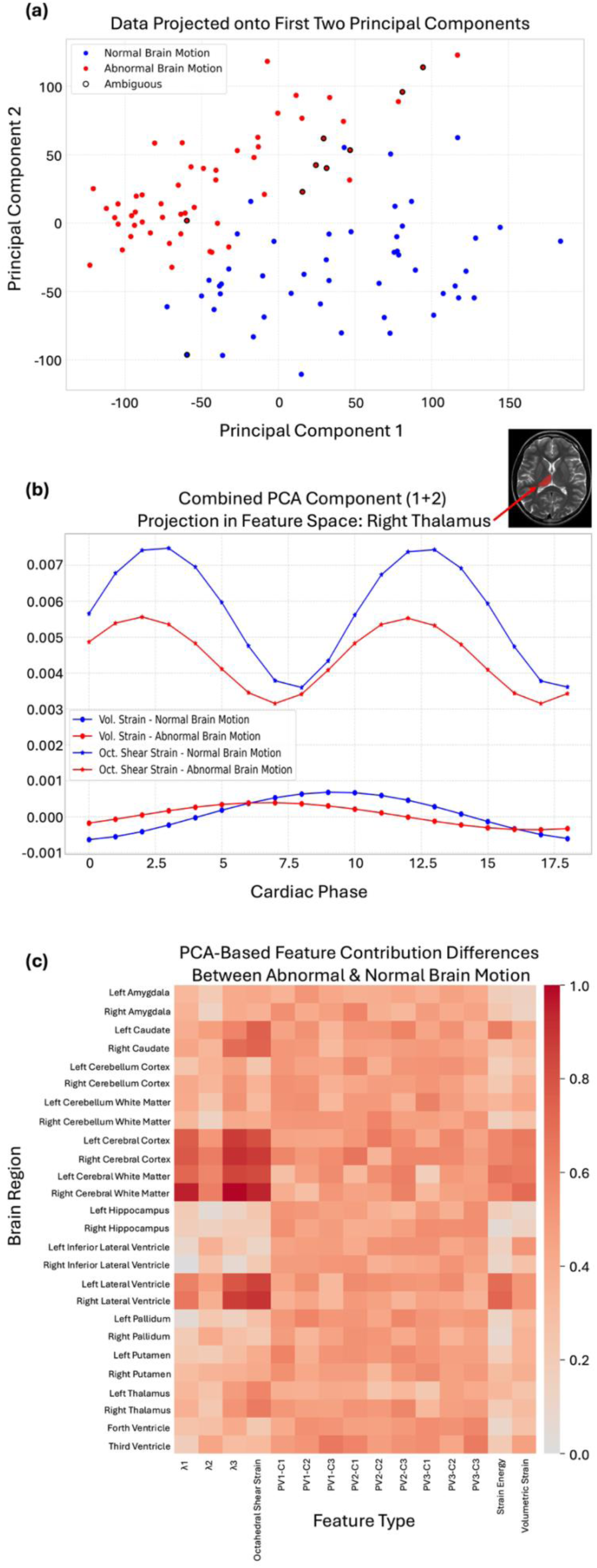
Principal component analysis of biomechanical features distinguishing normal and abnormal brain motion. (a) Projection of participant features onto the first two principal components, with normal and abnormal motion as labeled by five independent readers. Ambiguous consensus cases are highlighted with black circles and appear primarily near the separation boundary. (b) Sum of mean Principal Components 1 and 2 projected back into feature space across the cardiac cycle in the right thalamus, shown separately for the normal group (blue) and abnormal group (red). Two mechanical features are displayed: volumetric strain (Vol.) and octahedral shear strain (OCT). (c) Heatmap showing the contribution of PCA features to the differences between normal and abnormal motion by brain region and tensor-derived measures. PVX–CX represent the components of the principal vectors derived from the strain tensor, and λ1–λ3 represent the corresponding eigenvalues. Shown features were significantly different between groups after correction for multiple comparisons (FDR, p < 0.05).

To interpret these components in anatomical and mechanical terms, we reconstructed the significant principal components in feature space. The regions with the highest contributions included the cerebral white matter, cerebral cortex, lateral ventricles, caudate, and thalamus. Among tensor-derived biomechanical features, the most discriminative were octahedral shear strain, eigenvalues λ₁ and λ₃, strain energy, and volumetric strain. In terms of statistical descriptors, the mean and median contributed most strongly to group separation. Feature-level contributions were assessed for significance using permutation-derived empirical *p*-values with false discovery rate (FDR) correction (q < 0.05), ensuring that reported effects reflect robust group differences rather than chance.

Fig. 2b illustrates the summed group mean projection of PC1 and PC2 across the cardiac cycle in the right thalamus, comparing normal and abnormal groups for the mean volumetric strain and mean octahedral shear strain – two interpretable measures of tissue deformation. The abnormal group showed both reduced strain magnitude and temporal shifts relative to the normal group, suggesting altered timing and amplitude of cardiac-driven brain motion.

Feature-level contributions were summarized in a normalized heat map (Fig. 2c), organized by brain region and mechanical features. Strongest group discrimination was observed in the cerebral cortex and white matter for λ₁, λ₃, and octahedral shear strain, with similar though smaller effects in the lateral ventricles, caudate, and thalamus. Direction-dependent principal strain vectors contributed comparatively less than magnitude-based measures, indicating that differences between normal and abnormal motion are driven primarily by the amplitude rather than the orientation of tissue deformation.

Extended Data Fig. 1 and Fig. 2 further detail these findings. Extended Data Fig 1 (region × statistical descriptor) shows that mean, median, and variance contributed most strongly to group discrimination in the cortex, white matter, and ventricles. Extended Data Fig. 2 (region × cardiac phase) demonstrates that the lateral ventricles and caudate show peak separation around systole (phases 1–3, 18–19) and diastole (phases 8–12), whereas the cerebral cortex and white matter exhibit consistent group separation throughout the cardiac cycle, with pronounced peaks near both systole and diastole.

### Brain Motion Classification and Brain Age Estimation

To identify abnormal patterns of brain motion, we trained a logistic regression classifier using features derived from voxelwise strain tensors computed from displacement fields. Dimensionality reduction was first performed using PCA, retaining components that collectively explained 90% of the variance. The resulting principal components were used as inputs to a logistic regression model with L2 regularization and balanced class weights to account for class imbalance. The dataset was split into 79 participants for training and 26 participants for testing, and the procedure was repeated using stratified 4-fold cross-validation performed 25 times (100 independent train–test evaluations in total) to ensure robust performance estimates.

The classifier achieved high and consistent accuracy across repetitions. The average receiver operating characteristic (ROC) curve (Fig. 3a) yielded a mean area under the curve (AUC) of 99.4% ± 0.1% (95% CI). The overall confusion matrix (Fig. 3b) showed five misclassifications out of 105 participants – three false negatives and two false positives. Distributions of model performance across repeated runs (Fig. 3c) demonstrated strong reproducibility, with accuracy = 95.1% ± 0.7%, sensitivity = 95.4% ± 1.0%, specificity = 94.7% ± 1.2%, precision = 95.7% ± 1.0%, F1 score = 95.4% ± 0.6%, and AUC = 99.4% ± 0.1% (mean ± 95% CI).

**Fig. 3:**
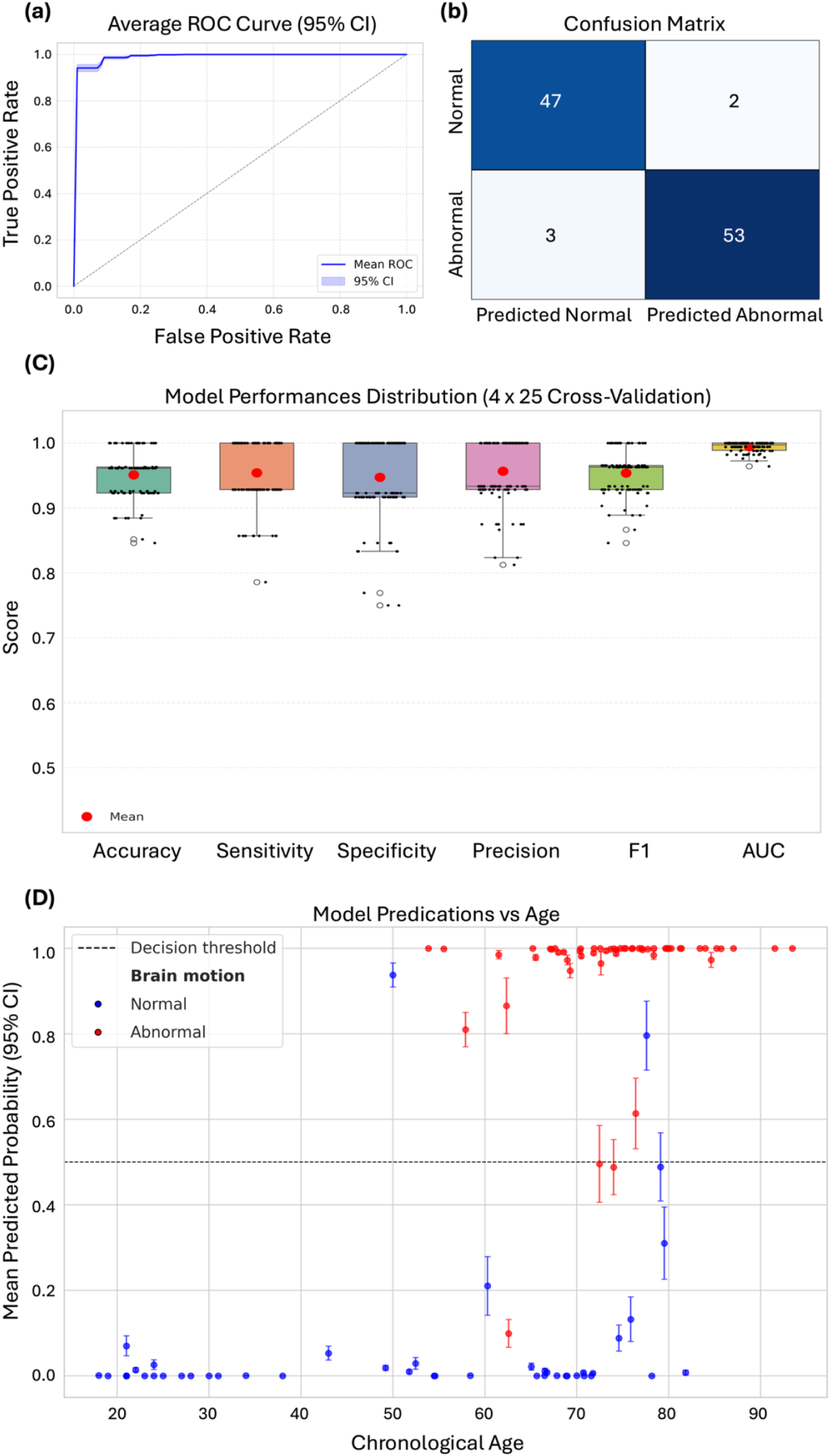
Logistic regression classification of abnormal versus normal brain motion. (a) Average ROC curve across 100 repeated runs (mean AUC = 99.4% ± 0.1%, 95% CI). (b) Confusion matrix comparing predicted labels to ground truth (5 misclassified cases: 3 false negatives, 2 false positives). (c) Distribution of model performance metrics (mean ± 95% CI): accuracy 95.1% ± 0.7%, sensitivity 95.4% ± 1.0%, specificity 94.7% ± 1.2%, precision 95.7% ± 1.0%, F1 score 95.4% ± 0.6%, AUC 99.4% ± 0.1%. Only 3 of 100 models had any metric below 80%. (d) Mean predicted probability (with 95% CI) of abnormal motion versus age, demonstrating a strong association between abnormal brain motion and increasing age.

Predicted probabilities of abnormal motion as a function of chronological age (Fig. 3d) revealed a strong age dependence, with misclassified cases highlighted above or below the decision threshold. Consistently, Extended Data Fig. 3 shows that several principal components were significantly correlated with chronological age (components 1–3: Pearson r = –0.456, 0.587, and 0.420; all *P* < 0.05), indicating that biomechanical strain features capture meaningful age-related variation.

Building on this observation, we next applied an Elastic Net regression model to estimate brain age from the same strain-derived features. PCA was performed on the training set (79 participants) and applied to the test set (26 participants), retaining components explaining 90% of the variance. The model was trained using a regularization parameter α = 0.01 and an L1 ratio of 0.5 to balance sparsity and smoothness.

Across 100 cross-validation iterations, the model achieved a mean absolute error (MAE) of 7.78 years (95% CI: ±0.23), a coefficient of determination (R²) of 0.756 (95% CI: ±0.017), and a Pearson correlation coefficient of 0.87 (p < 0.0001) between predicted and chronological age (Fig. 4a). The brain age gap (predicted age minus chronological age) showed a regression-to-the-mean pattern (Fig. 4b), with a significant negative correlation with chronological age (r = –0.64, *p* < 0.0001).

**Fig. 4:**
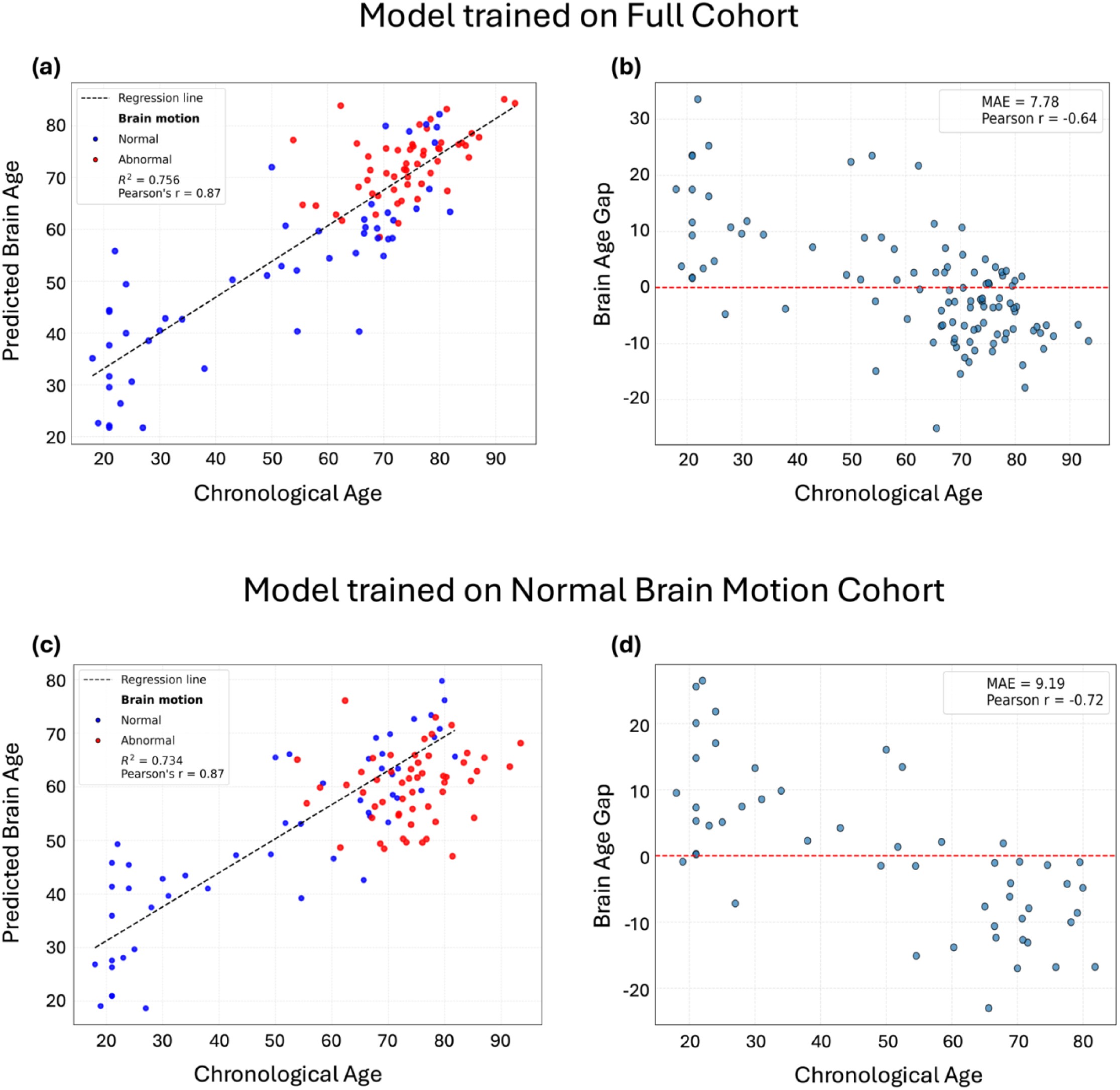
Elastic Net regression prediction of brain age from voxelwise strain tensor features. (a) Scatter plot of predicted versus chronological age for all participants. Individuals with normal motion are shown in blue, and those with abnormal motion in red. Model performance: MAE = 7.78 ± 0.23 years, R² = 0.756 ± 0.017, Pearson r = 0.87 (p < 0.0001). (b) Brain age gap (predicted minus chronological age) plotted against chronological age, demonstrating a regression-to-the-mean pattern (Pearson r = –0.64, p < 0.0001). (c) Scatter plot of predicted versus chronological age after training the model exclusively on participants with normal motion (n = 51, blue). Predictions for participants with abnormal motion (n = 54, red) are obtained by projecting them into the same model. Model performance: MAE = 9.19 years, R² = 0.734, Pearson r = 0.87 (p < 0.0001). (d) Brain age gap as a function of chronological age for the normal-motion test set, again showing a regression-to-the-mean effect (Pearson r = –0.72, p < 0.0001). Note that the brain age gap is computed only for the normal-motion participants used in model training, resulting in fewer data points compared to panel (b).

To minimize potential bias introduced by abnormal motion and to isolate typical motion–age relationships, we retrained the Elastic Net regression model using only participants with normal brain motion (n = 51). This model therefore captures age-related variation in biomechanical features under normative motion conditions. Participants with abnormal motion (n = 54) were then projected onto this model to evaluate how their estimated brain age diverged from the normative regression. The model maintained strong performance (R² = 0.734, r = 0.87, p < 0.0001, MAE = 9.19 years) (Fig. 4c), and many abnormal-motion participants showed substantial deviations from the regression line, indicating disrupted or atypical motion–age relationships. Within the normal-motion training cohort, the brain age gap again exhibited a regression-to-the-mean pattern (r = –0.72, p < 0.0001) (Fig. 4d).

Fig. 4c depicts the results with labels indicating only normal versus abnormal motion. In Fig. 6a, we show the same projections but with clinical diagnoses overlaid: normal-motion participants appear in blue and abnormal-motion participants in red, with distinct marker shapes denoting dementia subtypes. This visualization highlights that many clinically diagnosed participants fall within the abnormal-motion group and deviate markedly from the normative regression line, whereas some healthy controls also exhibit abnormal motion and similarly fall outside the expected age trajectory.

### PCA Features Analysis After Age Regression

Given that strain tensor-derived features in PCA space are strongly correlated with chronological age, we first selected participants older than 50 years (n = 78) and regressed out the effect of age. To identify biomechanical features that best differentiate normal from abnormal brain motion, we performed PCA on the reduced feature matrix and linearly regressed out the effect of age from the resulting components. The degree of group separation along each component was quantified using Cohen’s d. This analysis revealed that 20 principal components exceeded the conventional threshold for a small but meaningful effect size (|d| ≥ 0.2), with PC1 and PC2 showing the strongest separation (|d| = 0.95 and 1.35, respectively).

Projection of participants onto the first two components (Fig. 5a) revealed a clear separation between cases classified as normal and those with abnormal motion based on five-reader consensus. The separation was slightly reduced compared to the analysis prior to age regression, with a few normal and abnormal cases overlapping. Notably, participants for whom expert readers had difficulty reaching consensus (marked with black circles) clustered near the boundary between normal and abnormal groups, indicating that their motion patterns were also intermediate in the model’s feature space.

**Fig. 5:**
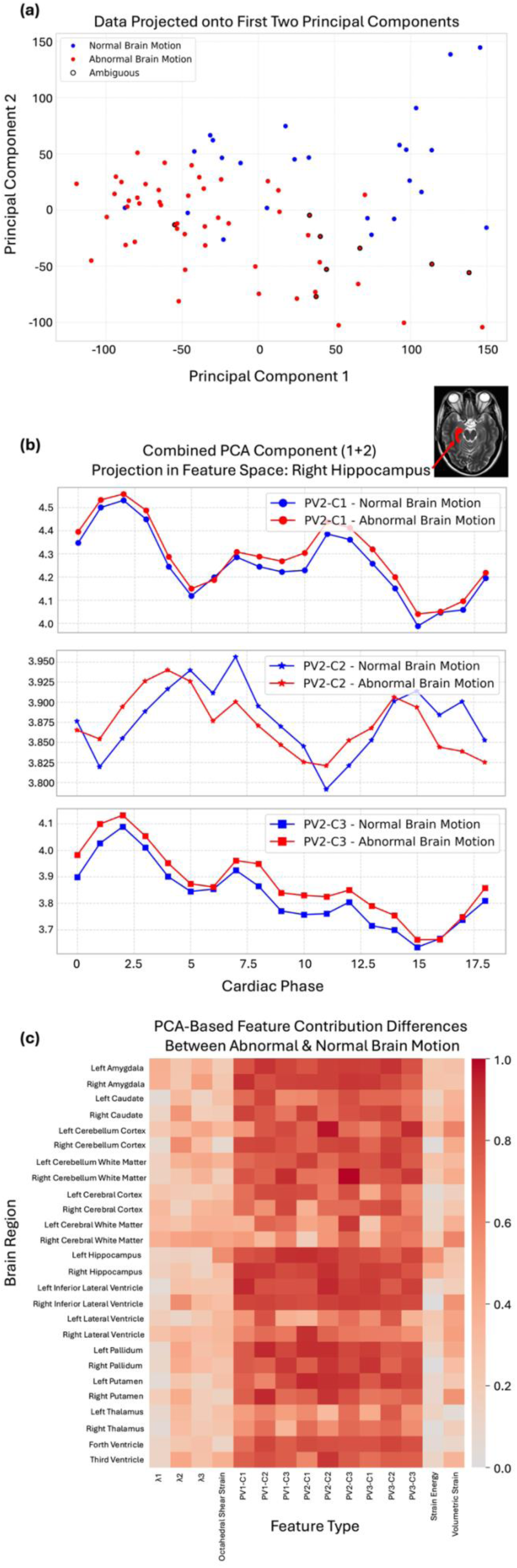
Principal component analysis of age-regressed biomechanical features in participants >50 years. (a) Projection of participant-level features onto the first two principal components. Normal and abnormal motion classifications are shown based on consensus from five independent readers; cases without clear consensus are outlined in black and cluster near the decision boundary. (b) Principal components 1 and 2 reconstructed in feature space across the cardiac cycle in the right hippocampus. Curves are shown separately for the normal (blue) and abnormal (red) groups for three tensor-derived biomechanical features: the three components (C1, C2, C3) of the second principal vector (PV2). (c) Heat map summarizing the contribution of PCA-derived features to group differences across brain regions and tensor-derived measures. PVX–CX denote components of principal vectors derived from the strain tensor, and λ1–λ3 denote the corresponding eigenvalues. Displayed features remained significantly different between groups after multiple-comparison correction (FDR, p < 0.05).

To interpret these components in anatomical and mechanical terms, we reconstructed the significant principal components in feature space. The highest contributing regions included the hippocampus, inferior lateral ventricles, amygdala, cerebellum, pallidum, putamen, and fourth ventricle. The dominant discriminative features were primarily associated with principal strain vectors, indicating that group differences were now driven mainly by the orientation of tissue deformation rather than its magnitude, which had explained most of the separation in the previous PCA analysis before age regression. Among statistical descriptors, kurtosis and skewness contributed most strongly to group separation. Feature-level contributions were tested for significance using permutation-derived empirical p-values with false discovery rate (FDR) correction (q < 0.05), ensuring that reported effects reflect robust group differences rather than chance.

Fig. 5b illustrates the sum group mean projection of PC1 and PC2 across the cardiac cycle in the right hippocampus for the kurtosis of the second principal vector. The abnormal group showed distinct waveform shapes compared to the normal group, suggesting altered motion direction throughout the cardiac-driven brain pulsation.

Feature-level contributions were summarized in a normalized heat map (Fig. 5c), organized by brain region and mechanical features. The strongest group discrimination was observed in the hippocampus, inferior lateral ventricles, amygdala, and cerebellum for direction-dependent principal strain vectors. In contrast, features reflecting deformation magnitude–such as eigenvalues, volumetric strain, octahedral shear strain, and strain energy–contributed substantially less after age correction, indicating that group differences were primarily driven by directional rather than amplitude-related motion properties.

Extended Data Fig. 4 and Fig. 5 further detail these findings. Extended Data Fig. 4 (region × statistical descriptor) shows that kurtosis and skewness contributed most strongly to group discrimination in the hippocampus, inferior lateral ventricles, amygdala, putamen, and fourth ventricle. Extended Data Fig. 5 (region × cardiac phase) demonstrates consistent group separation across the cardiac cycle for these regions.

### Association of Brain Motion with Clinical Diagnosis and Amyloid Status

We trained a logistic regression classifier to predict normal versus abnormal brain motion using age-regressed features derived from voxelwise strain tensors, focusing on participants older than 50 years (n = 78). Here, we report results from the age-regressed model, which used PCA for dimensionality reduction, retaining the first seven components to prevent overfitting. PCA was performed independently within each training set to avoid information leakage. Model performance was evaluated using stratified 4-fold cross-validation repeated 25 times, yielding 100 independent train–test splits for stable and unbiased performance estimates. The classifier achieved high accuracy (84.3% ± 1.6% [mean ± 95% CI]) with sensitivity = 84.9% ± 1.9%, specificity = 83.2% ± 3.0%, precision = 92.3% ± 1.3%, F1-score = 88.1% ± 1.3%, and AUC = 90.7% ± 1.8%.

As shown in Fig. 6b, the predicted probability of abnormal motion is plotted against chronological age for visualization purposes, with normal (blue) and abnormal (red) motion cases clearly separated; distinct marker shapes again denote dementia subtypes. Even after regressing out age prior to model training, most participants with a clinical diagnosis remained above the model’s decision threshold, indicating a strong correspondence between abnormal brain motion and clinical status. Fig. 6c summarizes predicted classifications across diagnostic categories. In this older cohort, while some healthy controls were classified as abnormal (18 normal, 16 abnormal), the majority of participants with a clinical diagnosis were classified as having abnormal motion (8 normal, 36 abnormal). Fisher’s exact test confirmed a significant association between dementia and abnormal motion (odds ratio = 5.06, 95% CI: 1.83–14.04, *p* = 0.0014), indicating that individuals with dementia are approximately five times more likely to exhibit abnormal brain motion than healthy controls.

**Fig. 6:**
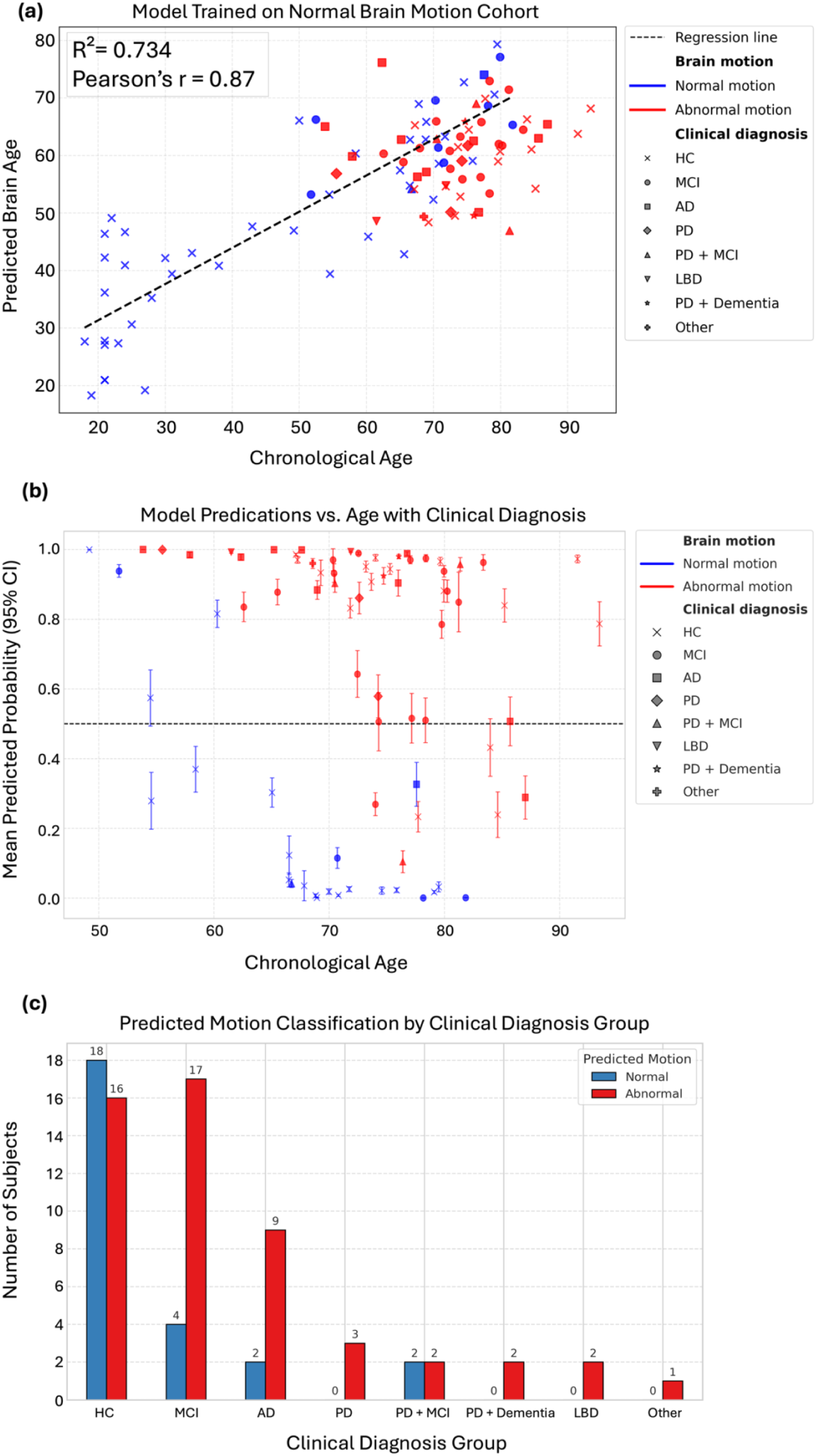
Association of brain motion with clinical diagnosis. (a) Scatter plot of predicted brain age versus chronological age using the same Elastic Net model shown in Fig. 4c, but here overlaid with clinical diagnosis markers. Participants with normal motion are shown in blue and those with abnormal motion in red; marker shapes denote dementia subtypes. The model achieved R² = 0.743, Pearson r = 0.87 (p < 0.0001), and MAE = 9.19 years. Some participants with a clinical diagnosis deviate from the estimated regression line. The pattern illustrates that many clinically diagnosed individuals cluster within the abnormal-motion group and show substantial departures from the normative age trajectory, while a subset of healthy controls also display abnormal motion and similarly fall outside the expected trend. (b) Predicted probability of abnormal motion versus age from a logistic regression classifier using age-regressed features in participants over 50 years, with 95% confidence intervals. Most dementia cases lie above the decision threshold. (c) Distribution of predicted motion classifications across clinical groups. While a notable fraction of healthy controls were classified as abnormal (18 normal, 16 abnormal), the majority of participants with a clinical diagnosis were classified as abnormal (8 normal, 36 abnormal). Fisher’s exact test shows a significant association between dementia and abnormal motion (odds ratio = 5.06, 95% CI: 1.83–14.04, p = 0.0014), indicating that dementia increases the likelihood of abnormal brain motion approximately fivefold.

We further examined the relationship between abnormal motion and amyloid pathology. As shown in Fig. 7a, most amyloid-positive participants fell above the abnormal-motion threshold, consistent with impaired biomechanical dynamics. Fig. 7b summarizes predicted classifications by amyloid status: while a subset of amyloid-negative participants were classified as abnormal (21 normal, 26 abnormal), the majority of amyloid-positive individuals exhibited abnormal motion (5 normal, 26 abnormal). Fisher’s exact test revealed a significant association between amyloid positivity and abnormal motion (odds ratio = 4.02, 95% CI: 1.38–12.83, *p* = 0.0077), suggesting that amyloid-positive individuals are about four times more likely to show abnormal motion than amyloid-negative individuals.

**Fig. 7:**
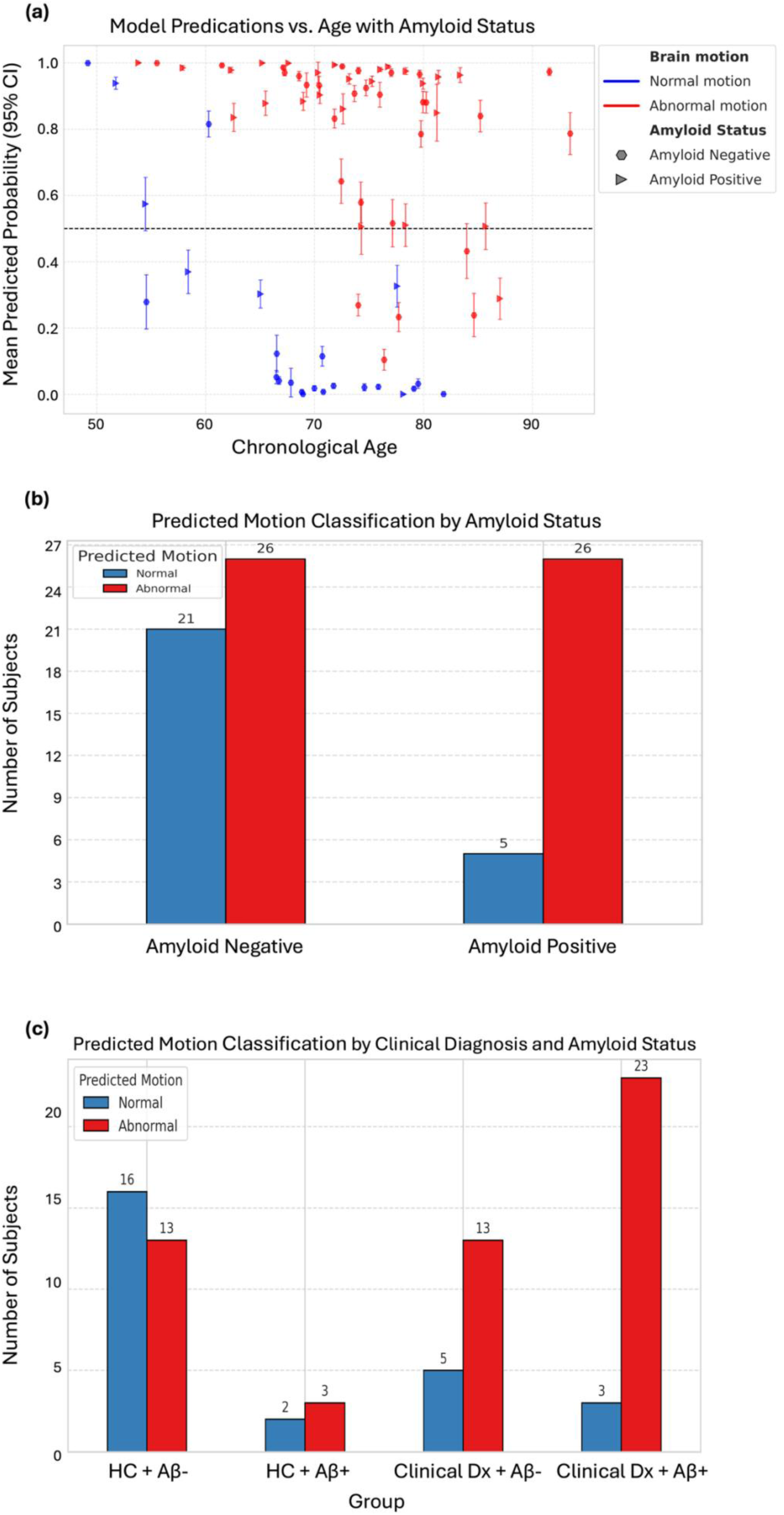
Association of brain motion with amyloid status. (a) Predicted probability of abnormal motion versus participant age, with 95% confidence intervals. Most amyloid-positive participants lie above the decision threshold. (b) Distribution of predicted motion classifications by amyloid status. While many amyloid-negative participants were classified as abnormal (21 normal, 26 abnormal), the majority of amyloid-positive participants exhibited abnormal motion (5 normal, 26 abnormal). Fisher’s exact test shows a significant association between amyloid positivity and abnormal motion (odds ratio = 4.02, 95% CI: 1.38–12.83, p = 0.0077), indicating that amyloid-positive individuals are approximately four times more likely to exhibit abnormal motion. (c) Predicted classifications stratified by combined clinical diagnosis and amyloid status (HC+AB–, HC+AB+, Diagnosis+AB–, Diagnosis+AB+). A substantial fraction of HC+AB– participants were classified as abnormal (13 abnormal vs. 16 normal), whereas the majority of Diagnosis+AB+ participants were abnormal (23 abnormal vs. 3 normal). Fisher’s exact test confirms a strong association for Diagnosis+AB+ participants (odds ratio = 9.44, 95% CI: 2.31–38.58, p = 0.0007), indicating nearly a tenfold increased likelihood of abnormal motion compared to amyloid-negative individuals.

Finally, Fig. 7c shows predicted classifications stratified by combined clinical diagnosis and amyloid status. Although a subset of healthy amyloid-negative participants were classified as abnormal (13 abnormal, 16 normal), the vast majority of participants with both a clinical diagnosis and amyloid positivity were abnormal (23 abnormal, 3 normal). Fisher’s exact test confirmed a strong association between clinical + amyloid-positive status and abnormal motion (odds ratio = 9.44, 95% CI: 1.86–24.65, *p* = 0.0023), indicating that individuals with both clinical diagnosis and amyloid positivity are nearly seven times more likely to exhibit abnormal motion than amyloid-negative participants.

Consensus labels assigned by expert readers were strongly associated with clinical diagnosis. Across the cohort, Fisher’s exact test confirmed a significant association between dementia and reader-classified abnormal motion (odds ratio = 7.12, 95% CI: 2.39–21.26, *p* = 0.0002), indicating that participants with a clinical diagnosis were roughly seven times more likely to be rated as having abnormal motion than healthy controls. The relationship between amyloid positivity and abnormal motion was weaker and did not reach conventional significance (odds ratio = 2.59, 95% CI: 0.89–7.52, *p* = 0.062), consistent with a trend toward higher prevalence of abnormal motion among amyloid-positive individuals. Importantly, participants with both a clinical diagnosis and amyloid positivity showed a strong association with abnormal motion (odds ratio = 8.21, 95% CI: 2.01–33.53, *p* = 0.0016), indicating that this combined subgroup is substantially more likely to exhibit abnormal motion than amyloid-negative participants.

## Discussion

In this work, we provide the first in vivo characterization of cardiac-driven whole-brain pulsatility and strain-based biomechanical features across human aging and dementia. Leveraging 3D q-aMRI, we reveal a clear age-related biomechanical signature of the brain. Even after rigorously controlling for age, individuals with dementia exhibit persistent and pronounced alterations in strain-derived metrics. These results indicate that cardiac-induced brain biomechanics – captured dynamically over the cardiac cycle – may function as an early and sensitive marker of disrupted neurophysiological homeostasis, offering new mechanistic insight into the onset and progression of neurodegeneration.

The reader-based classification demonstrated strong consistency, with inter-reader Kappa values of 0.65–0.68, indicating substantial agreement in distinguishing ‘normal’ from ‘abnormal’ brain motion. Intra-reader reliability further showed that these evaluations were stable and reproducible, supporting the conclusion that the observed motion differences reflect a genuine, visually discernible biomechanical pattern rather than subjective variability. Consensus discussions helped refine the operational criteria, emphasizing that under ‘normal’ motion, the ventricular system and midline structures should exhibit superior–inferior displacements of comparable magnitude. Incorporating these refinements, along with structured training, is expected to further improve inter-reader reliability.

Although the distinction between ‘normal’ and ‘abnormal’ brain motion remains an empirically defined construct, the strong convergence between expert ratings and data-driven PCA analysis suggests that it reflects an underlying biomechanical reality. Brain motion followed two separable distributions that expert readers could reliably distinguish, and PCA of strain-tensor features independently reproduced this separation without any knowledge of group labels. Given this clear separation in PCA space, it was unsurprising that the trained logistic regression model was able to distinguish ‘normal’ from ‘abnormal’ motion with high accuracy. Notably, ‘abnormal’ motion was observed predominantly in older individuals, consistent with prior reports of age-related reductions in brain stiffness and elasticity^15,16,46^.

At the same time, the strong age dependence of these features – and their ability to predict chronological age with high accuracy – indicates that this binary classification is likely a simplified abstraction. Rather than falling into two discrete categories, brain motion patterns appear to vary continuously across the lifespan. Indeed, the trained Elastic Net regression model demonstrated that cardiac-induced brain motion varies along a continuous spectrum rather than forming a strict binary division. This finding extends prior work showing age-related changes in the biomechanical properties of the brain^16,46,47^ and more recent evidence that brain volume, stiffness, and damping ratio collectively predict chronological age^48^. The brain-age scatterplot exhibited a characteristic regression-to-the-mean pattern, producing a mild negative bias, a well-known consequence of regression dilution^49^. Although this bias^50^ could be corrected, biological brain-age prediction was not the purpose of our study. Rather, our primary objective was to assess whether 3D q-aMRI yields biologically meaningful biomarkers. When trained exclusively on participants with normal motion, the model consistently identified individuals with abnormal motion as deviating from the expected aging trajectory. This reinforces the conclusion that strain features derived from 3D q-aMRI capture meaningful alterations in cardiac-driven brain biomechanics and may serve as early indicators of dysfunction. In addition, when clinical diagnoses were overlaid on these projections, many participants with neurodegenerative conditions fell within the abnormal-motion group and showed deviations from the normative trajectory. Conversely, some healthy controls also exhibited abnormal motion and were similarly displaced from the expected aging pattern. Together, these findings indicate that deviations from the biomechanical aging trajectory are not uniquely tied to clinical diagnosis. Although many participants with neurodegenerative conditions exhibit abnormal motion, a substantial fraction of abnormal-motion cases occur among cognitively normal controls. Thus, deviations in the biomechanical aging curve may arise from a mix of factors highlighting that abnormal motion alone cannot be interpreted as a definitive marker of pathology.

The multiple-comparison analysis performed before age regression indicated that the regions most responsible for distinguishing ‘normal’ from ‘abnormal’ brain motion were the lateral ventricles, cerebral white matter, cerebral cortex, caudate, and thalamus. These regions are well known to undergo structural changes during normal aging^51–53^, which may explain why they contributed strongly to the group separation. Notably, these regions were also identified by expert readers as key features distinguishing the two motion patterns. It is important to note, however, that the ventricular system is filled with CSF, so strain-based measures in these regions should not be interpreted as reflecting true tissue mechanical properties. Rather, the biomechanical features reflect the deformation and compliance of the ventricular shape as it responds to cardiac-driven pulsations.

After regressing out age, restricting the analysis to older participants (>50 years), and applying multiple-comparison correction, the regions driving group differences shifted to the hippocampus, inferior lateral ventricles, amygdala, cerebellum, pallidum, and putamen – most of which are well – established regions of involvement in dementia^54–56^, with the cerebellum only more recently recognized^57,58^. In this age-restricted analysis, the dominant mechanical features were the strain eigenvectors, capturing the direction of deformation. This is particularly notable because expert readers consistently highlighted two main characteristics of abnormal brain motion: reduced amplitude and asymmetry. By contrast, in the full cohort prior to age regression, the most discriminative features were those capturing overall deformation magnitude – specifically, the principal strain eigenvalues, octahedral shear strain, strain energy, and volumetric strain. Together, these findings suggest that reductions in overall brain motion primarily reflect normal aging, likely driven by arterial stiffening^59^, progressive decreases in cerebral stiffness^47^, and increased damping ratio^16^, whereas asymmetrical motion may be more associated with some pathological processes, though these interpretations are preliminary and warrant further rigorous investigation. We hypothesize that two physiological mechanisms may contribute to the observed asymmetry. First, because our method captures brain motion driven by cardiac-induced pulsatile blood flow, asymmetrical or delayed hemodynamics could lead to correspondingly asymmetric tissue deformation. Supporting this notion, recent study has reported that older adults exhibit significant interhemispheric differences in the cerebral pulsatility index, with higher values in the left hemisphere compared with the right, whereas younger adults show no such asymmetry^60^. Second, the asymmetrical motion may arise from structural or pathological hemispheric differences^61^. For example, a recent study demonstrated that hemispheric asymmetry in tau pathology is associated with asymmetric amyloid deposition in Alzheimer’s disease^62,63^. Together, these findings suggest that both vascular and structural factors may contribute to the asymmetric motion patterns detected by 3D q-aMRI.

Unlike prior MRE studies of aging and dementia^15,17^, which typically utilize an external actuator to induce dynamic shear waves and provide only a single, static snapshot of brain mechanics, our 3D q-aMRI approach measures natural, cardiac-driven deformation continuously across the entire cardiac cycle. This allows for the calculation of time-dependent strain tensor maps that capture how tissue biomechanics change with each heartbeat. While certain time-varying statistical descriptors, such as skewness and kurtosis, are less intuitive to interpret mechanically, mean values across brain regions provide clearer insight. For instance, before accounting for age-related confounding, the abnormal group exhibited consistently reduced magnitudes of both volumetric and shear strain compared with the normal group throughout the cardiac cycle. Moreover, the strongest separation between groups occurred around peak systole and peak diastole, the moments of maximal deformation. After regressing out age, however, this pronounced separation became more uniform across the entire cardiac cycle, suggesting that the motion asymmetry persists throughout the cycle rather than being limited to moments of peak deformation.

After regressing out age and focusing on subjects older than 50, the logistic regression model revealed a strong association between abnormal brain motion and clinical diagnoses within the AD and LBD spectra (odds ratio ∼ 5), amyloid positivity (odds ratio ∼ 4), and the combination of both clinical diagnosis and amyloid status (odds ratio ∼ 9). These findings align with prior studies showing that age-related softening of the brain can be further exacerbated by neurodegenerative diseases, including Alzheimer’s disease, multiple sclerosis, and Parkinson’s disease^17,64–67^.

Notably, several participants who were cognitively normal and/or amyloid-negative still exhibited abnormal brain motion. The underlying cause of this motion is unknown, but previous studies have reported altered brain dynamics in other neurological conditions, such as hydrocephalus^68,69^, and Chiari Malformation^70^ type 1. Furthermore, our analysis did not account for additional physiological or lifestyle variables, such as body mass index, exercise habits, or medications (e.g., cardiovascular drugs), which could influence the observed motion patterns. Importantly, this work represents an exploratory study, and to the best of our knowledge, it is the first to apply cardiac-induced brain motion analysis, quantified through both displacement and extensive strain estimations, to a relatively large cohort.

The mechanistic links between arterial pulsation, brain tissue deformation, CSF flow, and metabolic waste clearance remain incompletely understood^10,71,72^. Although 3D q-aMRI captures only one component of this system, it provides a crucial missing piece: a direct, spatially resolved measure of how cardiac forces propagate through brain tissue. By mapping pulsation-driven strain throughout the entire brain, our method offers a mechanistic framework for probing how vascular pulsatility may couple to CSF dynamics and, ultimately, to clearance pathways.

Several limitations should be acknowledged. First, our analysis focused exclusively on motion at the fundamental cardiac harmonic, and we applied spatial smoothing to improve the signal-to-noise ratio of the displacement estimates. Although these steps enhanced robustness, they likely reduced sensitivity to higher-order harmonics and fine-scale motion, potentially obscuring additional physiological information. Future methodological developments may allow simultaneous noise suppression and recovery of richer motion dynamics, enabling more detailed subgroup analyses. Second, the relatively modest sample size, both among cognitively healthy controls and among participants with clinical diagnoses, together with the fact that most data were acquired on a single scanner, limits the generalizability of our results. Larger, multi-site studies with broader diagnostic representation will be essential for validation and refinement. Finally, regressing out age across the lifespan is nontrivial, given the complex and potentially nonlinear dependence of brain motion on age, particularly when younger adults are included. Such corrections risk either over-adjusting or masking the true disease-brain motion related effects.

In summary, this study demonstrates that 3D q-aMRI can quantify subtle cardiac-driven brain motion across the adult lifespan and that strain-based biomechanical features derived from these motions provide interpretable signatures of underlying tissue dynamics. Abnormal motion patterns showed broad associations with age, clinical diagnosis, and amyloid status, indicating that cardiac-driven biomechanics reflect both normative aging processes and disease-related alterations. At the same time, the presence of abnormal motion in some cognitively normal individuals underscores that biomechanical deviations are not tied to pathology in a one-to-one manner and likely reflect a combination of vascular, structural, and individual factors. Taken together, these findings establish brain biomechanics as an emerging dimension of brain health, with 3D q-aMRI offering a scalable, non-invasive modality for quantifying these dynamics in vivo. By providing a window onto the mechanical consequences of vascular and neurodegenerative processes, 3D q-aMRI may enable earlier detection of biomechanical vulnerability than is possible with conventional structural imaging.

## Data Availability

All data produced in the present study are available upon reasonable request to the authors

## Acknowledgment

The authors acknowledge all Stanford ADRC research participants and their families for their invaluable contributions. This work was supported by the National Science Foundation Graduate Fellowship (Grant No. 1828993); NIH grants R01MH116173, R01EB019437, U01EB025162, P41EB030006, and R21NS111415; the Royal Society of New Zealand Marsden Fund; and the Kānoa - Regional Economic Development & Investment Unit, New Zealand. We are grateful to our research participants for dedicating their time to this study. We also acknowledge the support of GE Healthcare. Additional funding for this work was provided by the National Institutes of Health Stanford ADRC (P30 AG066515), NIH/NIA 1K23AG090733-01A1 (KY), the Alzheimer’s Association AACSF-24-1307411, the Iqbal Farrukh and Asad Jamal Good Planet Foundation Fund, and the Fred and Barbara Erb Family Foundation.

## Extended Data

**Extended Data Fig. 1.**
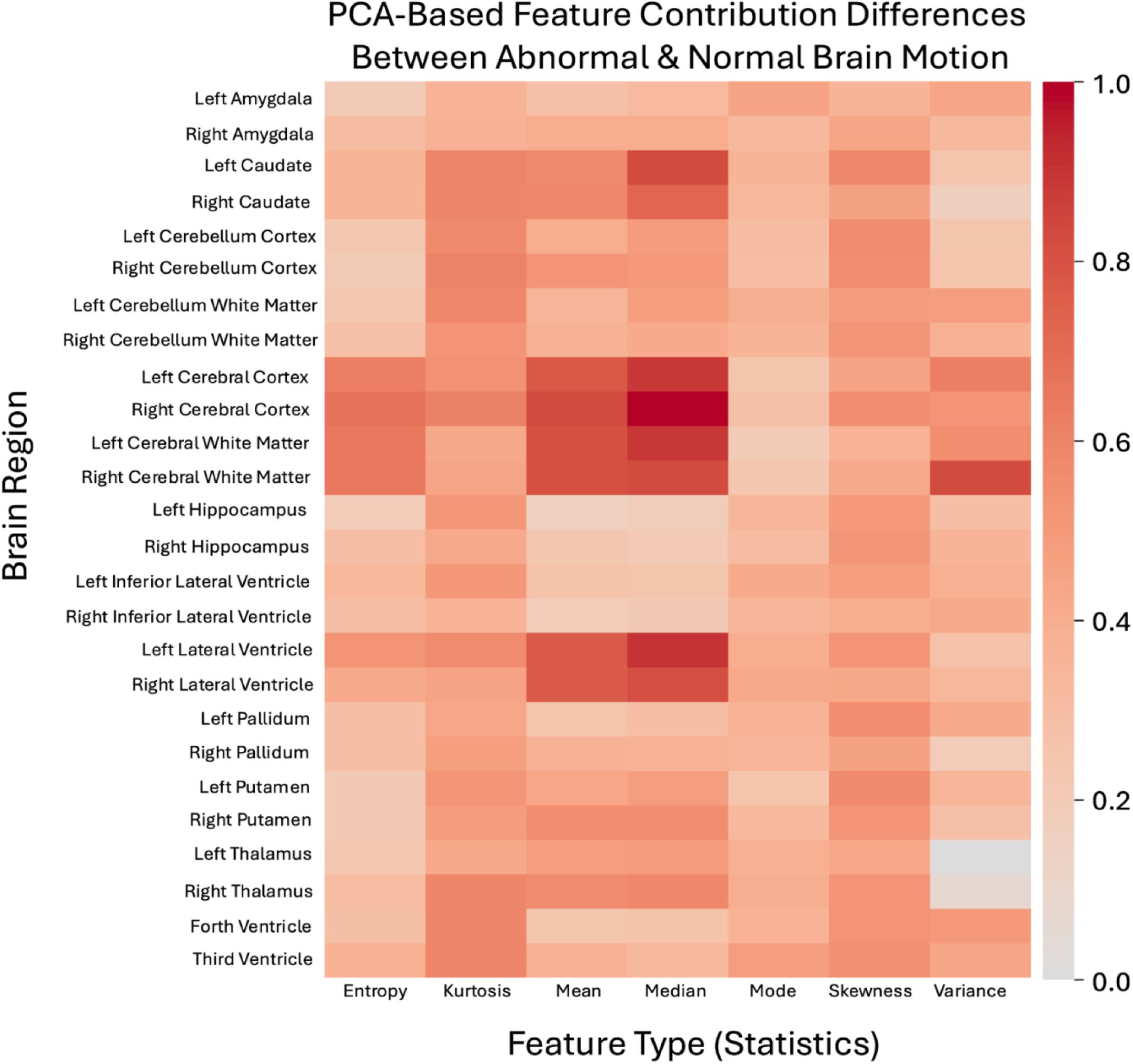
Heatmap of PCA feature-contribution differences between abnormal and normal brain-motion groups prior to age regression, organized by tensor-derived biomechanical measures and statistical descriptors. The strongest contributions arise from the cerebral cortex, white matter, and lateral ventricles, particularly for mean and median descriptors.

**Extended Data Fig. 2.**
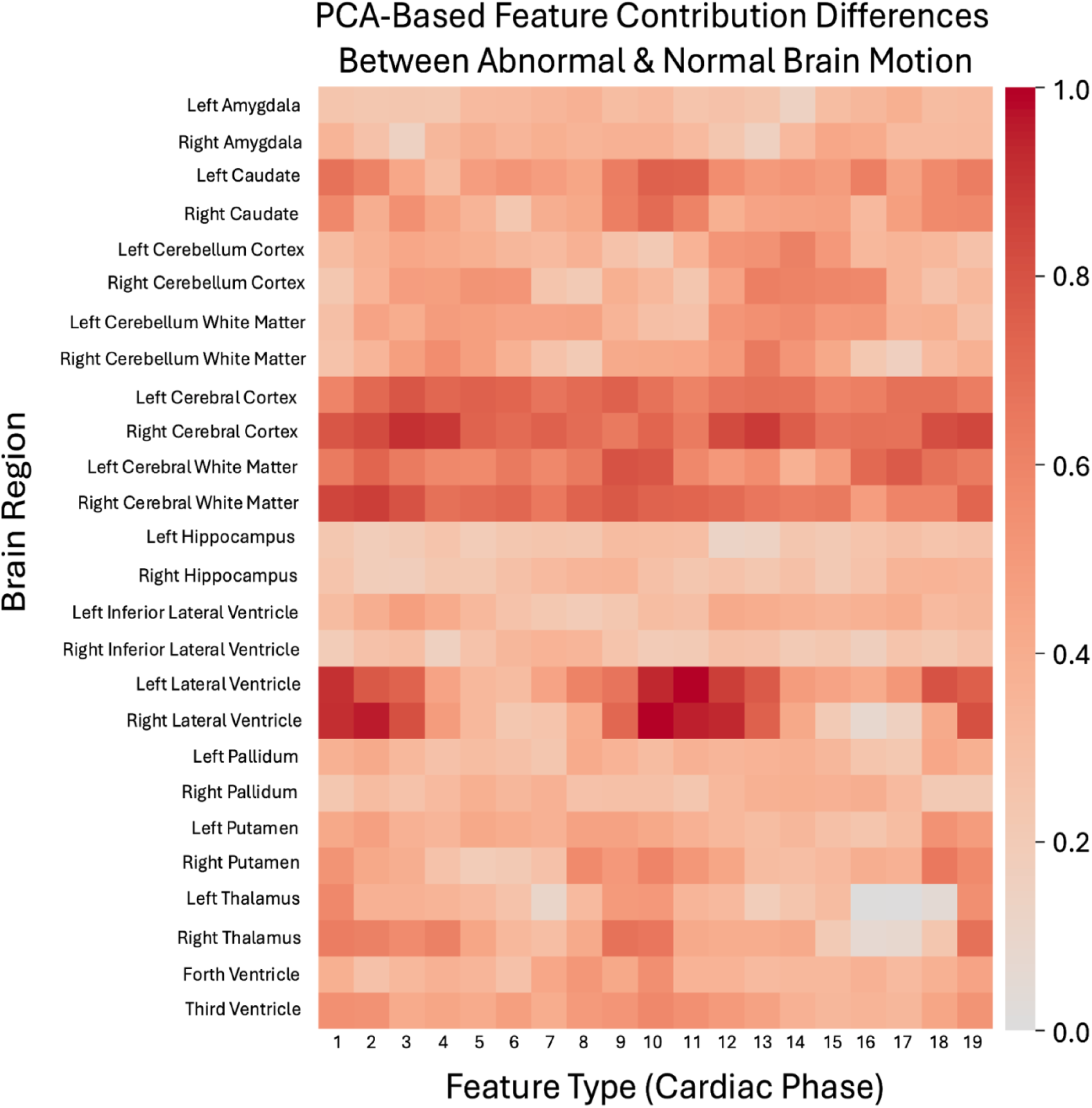
Heatmap of PCA feature-contribution differences between abnormal and normal brain-motion groups prior to age regression, organized by brain region and cardiac phase. The cerebral cortex and cerebral white matter show strong separation across the full cardiac cycle, whereas the lateral ventricles exhibit separation predominantly near peak systole and diastole.

**Extended Data Fig. 3.**
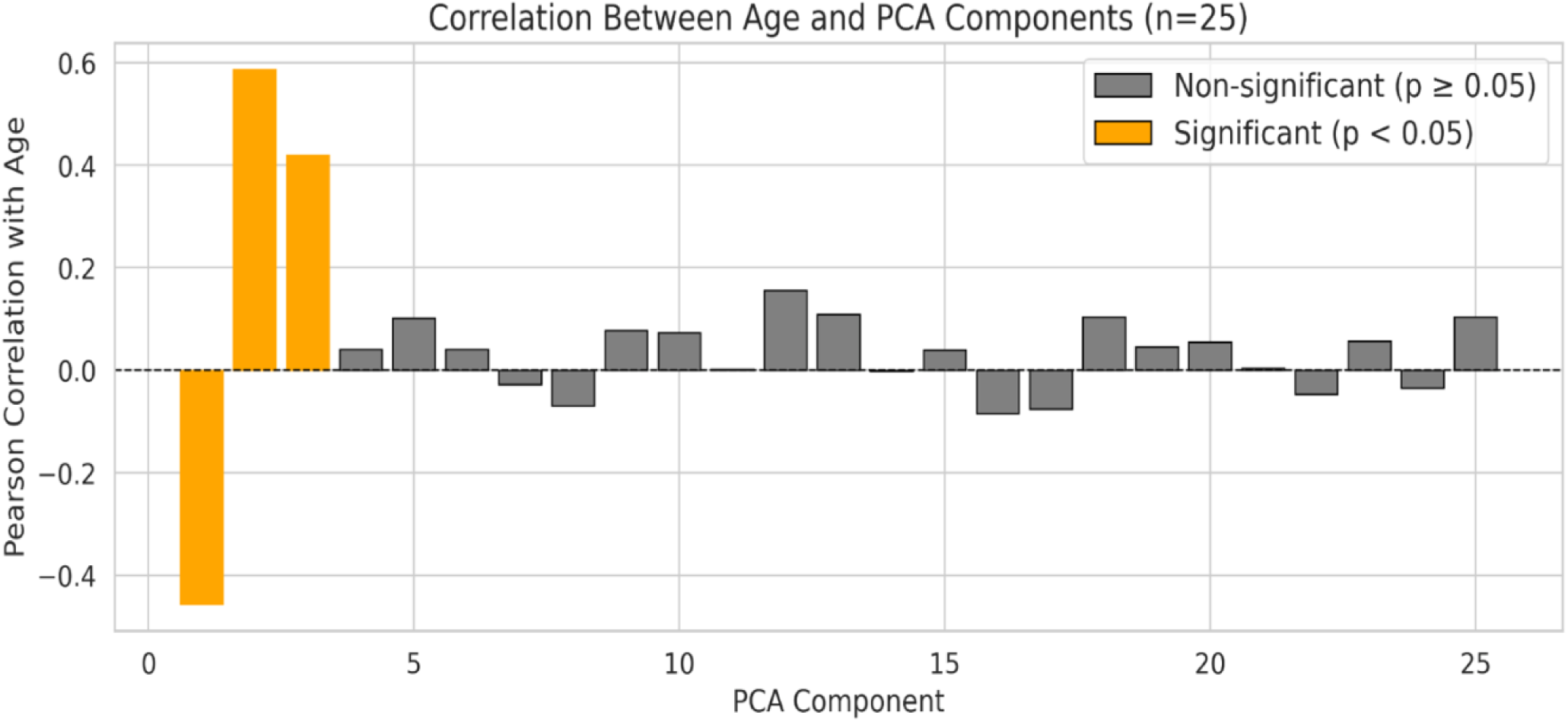
Correlation between the first 25 PCA components (derived from the full dataset) and chronological age. Components 1–3 show significant associations with age (r = –0.456, 0.587, and 0.420; P < 0.05), demonstrating a strong relationship between strain-derived biomechanical features and aging.

**Extended Data Fig. 4.**
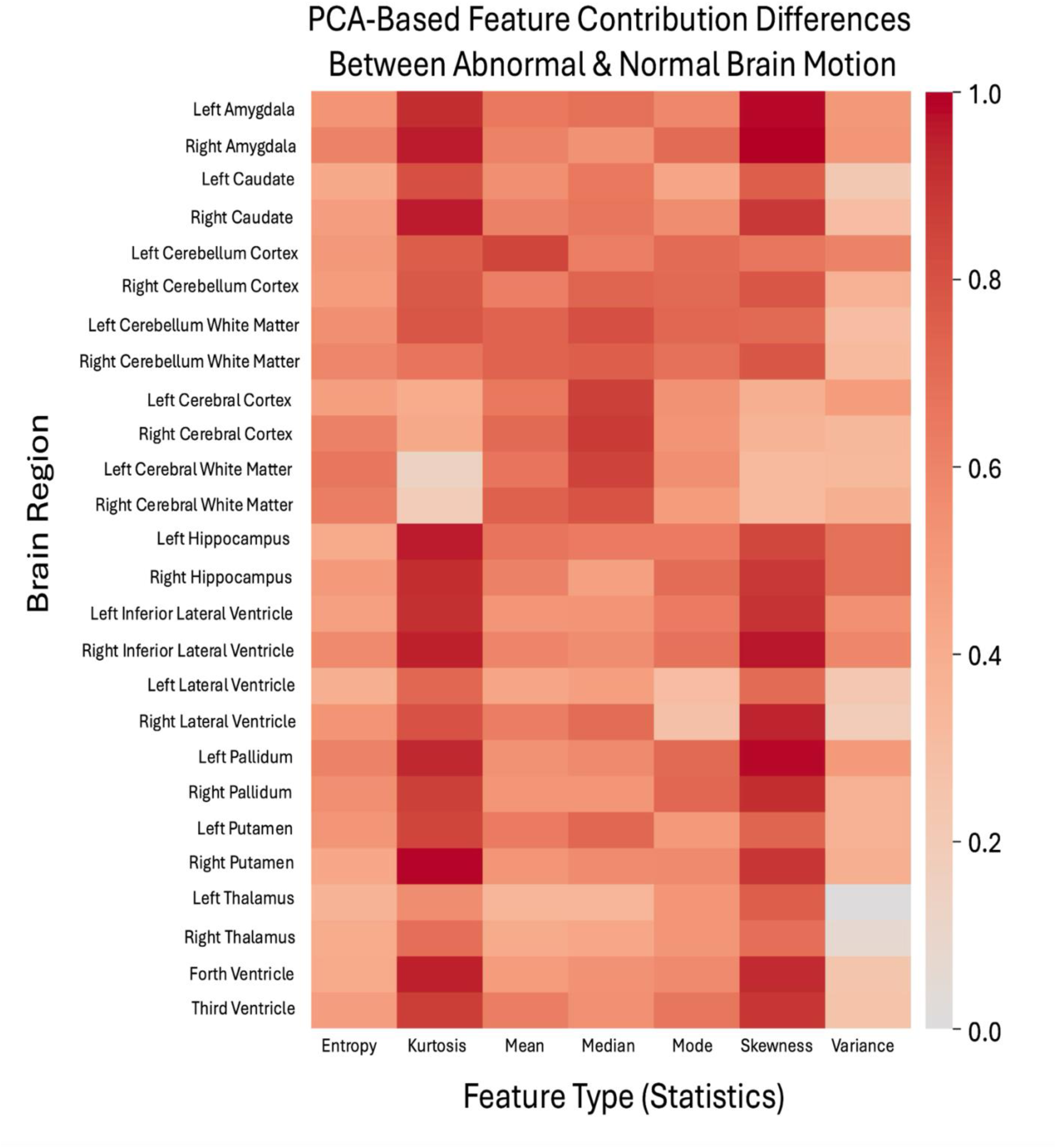
Heatmap of PCA feature-contribution differences between abnormal and normal brain-motion groups after age regression, organized by tensor-derived biomechanical measures and statistical descriptors. The strongest contributions arise from the hippocampus, amygdala, inferior lateral ventricles, putamen, and fourth ventricle, with particularly pronounced effects in kurtosis and skewness.

**Extended Data Fig. 5.**
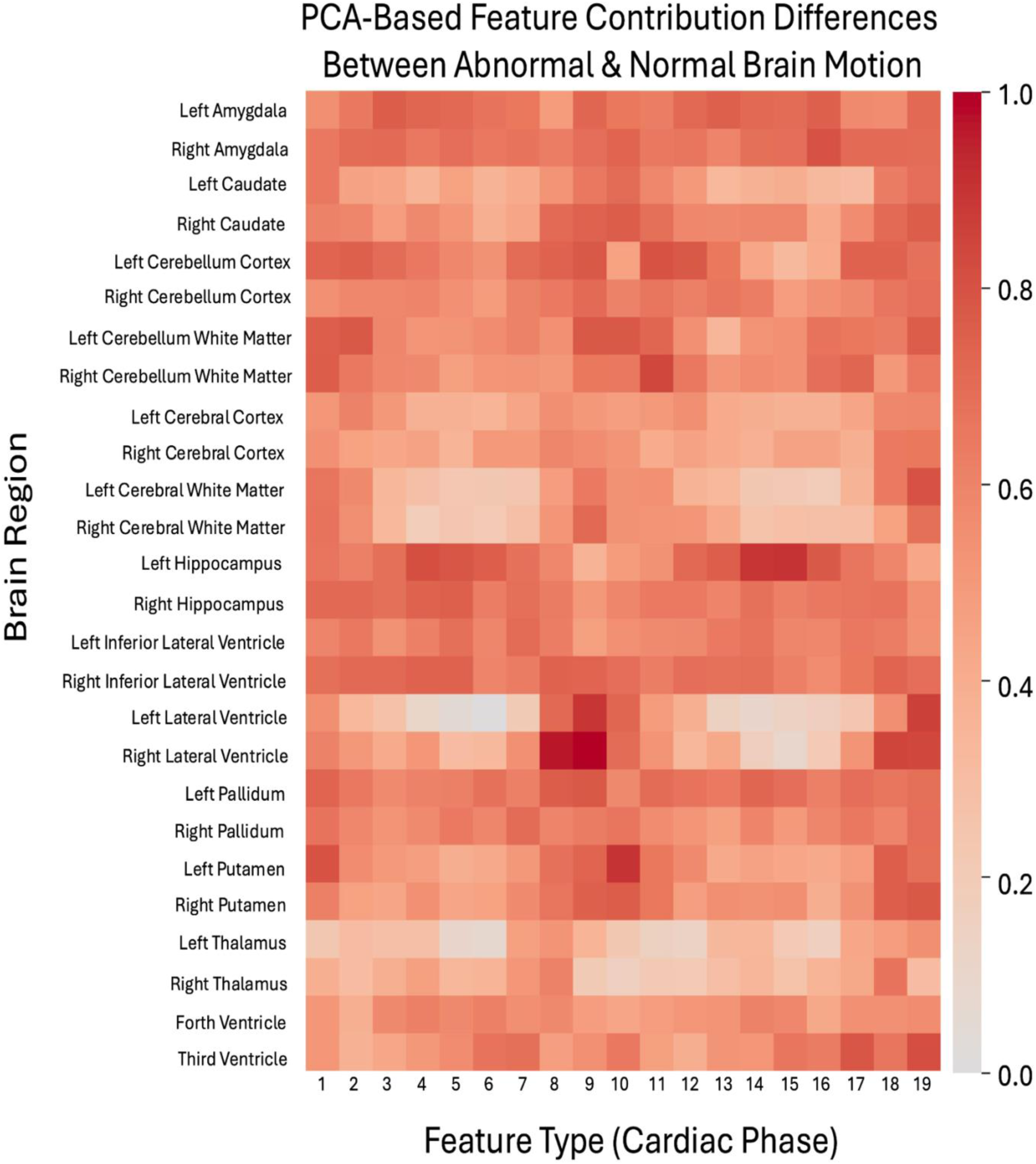
Heatmap of PCA feature-contribution differences between abnormal and normal brain-motion groups after age regression, organized by brain region and cardiac phase. The hippocampus, amygdala, and inferior lateral ventricles show strong separation across the full cardiac cycle.

## Abbreviations

AD: Alzheimer’s disease
CSF: cerebrospinal fluid
LBD: Lewy body disease
3D q-aMRI: 3D quantitative-amplified MRI

